# Building comprehension to enhance vaccination intentions: Evidence from the United States and Italy

**DOI:** 10.1101/2020.06.05.20122986

**Authors:** Greg Porumbescu, Maria Cucciniello, Paolo Pin, Alessia Melegaro

## Abstract

Building on Cognitive Load Theory and the Elaboration Likelihood Model of Persuasion this study attempts to deepen our understanding of how health information campaigns related to vaccination work. We predict that including statistical information in health information campaigns will serve as a distraction that suppresses comprehension that renders the campaign less effective at persuading the public to vaccinate. Results from a survey experiment conducted on samples of US (n = 605) and Italian parents (n = 505) show support for the hypothesized relationship depends upon the form of comprehension (comprehension of how vaccines work versus comprehension of potential adverse effects of vaccinating). Specifically, both US and Italian parents show including statistical information reduces comprehension of how vaccines work, in turn reducing parents’ intentions to vaccinate themselves and their children. We find no support for the hypothesized mechanism when comprehension of potential adverse effects of vaccinating acts as the process variable.

---

To strengthen our theoretical understanding of how health information campaigns work we build upon insight from cognitive load theory (CLT), which explains how the way facts are presented influence their comprehensibility (Sweller 1994; 2011), and the Elaboration Likelihood Model of Persuasion (ELM), which provides a foundation for understanding the cognitive processes that underly persuasion (Petty and Cacioppo 1986). Informed by these theoretical frameworks, we argue that the success of health information campaigns lies in the way they convey facts to the public. Specifically, we predict that using statistical information to convey facts to the public will serve as a distraction that suppresses comprehension and, in turn, renders the campaign less effective at persuading the public to vaccinate. To test the proposed mechanism, we design a survey experiment that explains how vaccines work as well as potential adverse effects of vaccinating, and then asks samples of parents from the United States (n = 605) and Italy (n = 505) to vaccinate themselves and their children. As we explain later in greater detail, we focus on vaccination compliance as an empirical case in light of data revealing that compliance rates for vaccination have been steadily declining over the past decade (World Health Organization 2017).

Results across samples of US and Italian parents turn back two key findings. First, they identify a specific form of comprehension – process comprehension – as a critical and distinctive mechanism responsible for translating health information campaigns into citizen compliance. Second, we demonstrate how the way health information campaigns communicate facts – using statistical information versus nonnumeric descriptors – shapes process comprehension (i.e., how vaccines work), but not comprehension of possible implications of complying (i.e., adverse effects of vaccinating). These findings break new ground by demonstrating how facts can be communicated to prompt specific forms of comprehension that play a critical role in persuading members of the public to vaccinate

## Vaccination intentions and health information campaigns

Health information campaigns differ from other forms of government communication, such as performance information disclosure in that they are crafted with the explicit intention of communicating facts to elicit a specific type of behavior (e.g., vaccinating). For example, every fall and winter, public health departments disclose facts the government has collected about the flu and ask the public to vaccinate against influenza to reduce instances of the flu. The hope is that exposing the public to these facts will persuade them to engage in a pre-determined health related activity, such as vaccinating. The success of health information campaigns, in their current form, is determined by their ability to convey facts to the public in ways that convince them that engaging in some health behavior is important for them and potentially for others (Kopfman and Ruth-McSwain, 2011).

## When do Facts Persuade?

One of the most frequently used frameworks for understanding persuasion is the elaboration likelihood model (ELM) of persuasion (Cacioppo and Petty 1984; Petty and Cacioppo 1986). This framework is widely used in research on consumer behavior, public health, and mass media, though it is seldom used to understand the effects of government communications (Alon-Barkat 2019). The ELM argues that persuasion occurs via an elaboration continuum (Petty and Cacioppo 1986). At the lower end of the continuum, persuasion primarily results from an intuitive response to information and little mental effort is exerted – this is referred to as the peripheral path. Here, persuasion is influenced by features of the information (i.e., peripheral cues) that have no substantive contribution to the quality of information at hand, such as the source of the information or the amount of information (Chaiken, Liberman, and Eagly 1989). At the other end of the continuum, where elaboration is higher and greater mental effort exerted, persuasion results from one’s comprehension of the information – this is referred to as the central path to persuasion. Persuasion that occurs via the central path is influenced by the quality of logic of the information a decision-maker is presented with. Persuasion that primarily results from comprehension of information has a more enduring impact on the resulting attitude (Petty and Binol 2009). Therefore, within the context of health information campaigns persuasion that occurs via the central route is preferable to persuasion that occurs via the peripheral path. For this reason, our focus is on the central path to persuasion.

To trigger the central path to persuasion, an argument must be perceived as strong, and also understood (Cacioppo and Petty 1984). While prior ELM research has assessed the role of argument strength in persuading (for a review see Petty and Wegener 1998), this project differs in that our focus is on argument comprehension. One factor that influences ability to process and comprehend information is the presence of distractions (Petty and Cacioppo 1986). This is because, distraction weakens the impact of the information itself (Chaiken 1980; Petty and Brinol 2010). While distractions can be part of an individual’s environment (e.g., loud noises), they can also be embedded in a message. For example, Alon-Barkat (2019) shows that embedding symbols in public communications campaigns discourages the central path to persuasion by leading individuals to pay less attention to content.

In addition to the use of imagery, CLT shows the way facts themselves are described can act as a distraction that reduces information comprehension (Chandler and Sweller 1994). CLT is a prominent theoretical framework used by education psychologists to explain factors and processes that influence individuals’ ability to learn new information. Central to CLT is the concept of cognitive load – the amount of mental effort an individual must exert to process and comprehend facts and information. CLT shows that some ways of presenting facts can increase the amount of mental effort needed to process them and render them distractions, whereas others can enhance comprehension by reducing the mental effort needed to process them and, by the logic of the ELM trigger the central path to persuasion. A key insight from CLT for research on health information campaign effectiveness is that for facts to have an enduring impact on citizen compliance, they must be presented in ways that are easy to comprehend. As we explain in greater detail below, one way of doing so is to limit reliance on statistical information when conveying facts to the public.

## Hypotheses: Shaping compliance intentions through comprehension

### Hypothesis 1: When do Facts Shape Comprehension?

The question of how to effectively inform citizens is the subject of longstanding debate in the social sciences. For some time the underlying logic that has dominated such discussions is that comprehension stems from exposure to content – exposing the public to more information will, on average, make them better informed. Yet advances in information and communications technology, which allow governments to disseminate vast amounts of information to the public in real-time, have quickly revealed limits of this logic. Instead, in keeping with insight from CLT, growing emphasis is placed upon communicating information to the public in ways that are easy to process and comprehend.

One way of doing so is to avoid the use of numbers and statistical information when communicating facts (Clark, Ngyuyen, and Sweller 2011). For example, the Security and Exchange Commission’s Plain English Rule (421(d)) (1998) and the Obama Administration’s Plain Writing Act (2010) both recommend heavy reliance on statistical facts to communicate information to the public. Moreover, the Center for Disease Control’s *Vaccines and Preventable Diseases* webpage is oriented toward the use of nonnumeric as opposed to numeric descriptors in describing health trends to the public (Center for Disease Control 2018). The reasoning underlying CLT and these initiatives is that using statistical information to describe facts increases the mental effort individuals exert to process a message, meaning messages with statistics and numeric descriptors are more cognitively burdensome to process than functionally equivalent messages that forego the use of numbers (Sweller 1994).

In part this greater mental effort results from a tacit calculation that associates a numerical statistic with a corresponding non-numeric descriptor (i.e., 0.002% corresponds to very small). Because of the additional mental effort, mental effort increases and comprehension decreases. Indeed, one study by the United States Department of Education found roughly half of the United States population in 2002 had a difficult time in understanding even basic numerical information (Kirsch et al. 2002). Similar findings are also present in recent public management research that assesses how different methods of presentation shape comprehension of government information (Alon-Barkat 2019; Isett and Hicks 2018; Olsen 2017). In general, this nascent line of work reveals that statistical information tends to be more difficult for individuals to recall and ultimately comprehend (Olsen 2017; Porumbescu et al. 2017).

All told, existing theory and evidence leads to the expectation that the use of statistical information by health information campaigns to communicate facts to the public will, on average, impede comprehension.

*H1: The use of statistical information in health information campaigns will, on average, reduce individual comprehension of health information campaign messages*

*H2: From comprehension to intentions to vaccinate*

A wealth of research has highlighted the irrational underpinnings of individual decision-making and behavior, uncovering now well-known cognitive biases such as framing effects (Gallagher and Updegraff, 2011; Druckman, 2001) default bias (Davidai at al.,2012), and base-rate fallacies (Barbey and Sloman, 2007), as well as emotional considerations (Schwarz 2011). Yet, as our awareness of cognitive and emotional biases has grown over time, just how rational citizens are remains unclear (for recent contributions to this debate see: Achen and Bartels 2016; Ashworth, Bueno de Mesquita, and Friedenberg 2018).

ELM posits that argument comprehension is an important determinant of enduring persuasion, with downstream relevance contingent upon the quality of argument an individual is exposed to (Briñol and Petty 2006; Petty, Wells, and Brock 1976). That is, lasting shifts in attitudes and behaviors are predicated upon argument comprehension. Available evidence appears to align with this point, suggesting that, holding argument strength constant citizens will update their beliefs and attitudes in the direction of the information they are exposed to (Guess and Coppock 2018). For example, Cook, Jacobs, and Kim (2010) found that efforts to increase citizens’ confidence in social security by exposing them to information explaining how social security works significantly increased confidence in the policy. Complementing the work by Cook and colleagues, Boeri and Tabellini (2012) demonstrate that exposure to policy information can translate into more positive attitudes toward a policy, even when individuals dislike the policy in question. Coppack, Ekins, and Kirby (2018) uncover similar effects within the context of op-ed pieces. Here, the authors find that, despite large initial differences in opinions on target issues, readers of op-ed pieces pertaining to these issues tended to be persuaded by the arguments these pieces lay to bear, irrespective of party identification or socio-economic status. Moreover, Grigorieff and colleagues (2016) demonstrate that changes to an individual’s understanding of an issue, attributable to new information not only influence their attitudes, but also their behavioral intentions. Specifically, they show that exposing individuals to information about immigration increases their proclivity to donate to charities that support immigrant causes. All told, the greater the impact new information has on individuals’ understanding of a particular issue, the more likely this information is to change their attitudes and behaviors related to the issue.

Research on health literacy has also demonstrated linkage between comprehension and patient compliance. For example, one recent meta-analysis examined 220 articles assessing the relationship between health literacy – a basic understanding of information related to a health issue – published between 1948 and 2012, showing that higher levels of health literacy were associated with adherence to a range of physician prescribed behaviors (Miller 2016). Similarly, research by Jacobsen and colleagues (1999) has shown that prompting individuals with low levels of health literacy to learn about the pneumococcal vaccine significantly increased their compliance with requests to vaccinate.

Taken together, extant evidence leads to the expectation that comprehension of health information campaigns requesting individuals engage in some activity will be associated with stronger intentions to comply with this request.

*H2: On average, levels of comprehension of the information embedded in a health information will be positively associated with intentions to vaccinate*

## Empirical Setting

We explore these hypotheses with reference to intentions to comply with requests to vaccinate. Vaccination as a form of government compliance is relevant in that data reveals this form of compliance has steadily decreased over the past decade in Europe (Larson et al. 2018) and the United States (Nadeau 2015). The decline in compliance with requests to vaccinate have been accompanied by increases in the frequency and severity of outbreaks of infectious diseases, such as the measles (World Health Organization 2017). In particular, Italy and the United States have experienced a dramatic increase in the number of measles outbreaks in recent years. As such, this form of compliance is particularly salient.

A recent report by the European Commission on Disease Prevention and Control (European Centre for Disease Prevention and Control 2017) shows health information campaigns seeking to combat vaccination hesitancy and enhance compliance requests to vaccinate tend to focus on bolstering public awareness of how vaccinations work and the implications and risks of vaccinating. However, extant trends suggest that the way these campaigns convey facts to the public have so far been ineffective in persuading members of the public to comply with requests to vaccinate. To this end, a second reason the vaccination setting is relevant to the purposes of this study is that available evidence suggests current approaches to using health information campaigns to increase compliance with requests to vaccinate are ineffective.

## Research Design

We use a randomized survey experiment to examine the causal impact of statistical information on health information campaign comprehension and vaccination attitudes for the following reasons. Information in the prompts was not altered in order to keep the strength of the argument constant. First, written health information campaigns are frequently used by governments to inform the public of the importance of getting vaccinated and to keep the abreast of various health risks confronting their community. Second, while a field experiment might provide causal estimates in real-life settings (greater external validity), the greater control afforded to researchers by a survey experiment permits a simpler, less noisy approach (greater internal validity) to estimating the causal effect of interest. That being said, a logical next step would be to examine our findings in a real life setting through the use of a field experiment or other methods.

We run our experiment using samples of parents from Italy and the United States because these contexts have different healthcare systems, political systems, and religious orientations, yet are both experiencing outbreaks of vaccine preventable infectious diseases and increases in vaccination hesitancy. As prior research on vaccination attitudes has shown, these dimensions are salient to the purposes of this study as they all are said to weigh on attitudes toward vaccination (Holmberg, Blume, and Greenough 2017). As such, by exploring our hypotheses across these distinctive contexts we are better able to explore the role statistical information and comprehension plays in influencing compliance intentions.

Additionally, we focus on parents’ attitudes toward vaccination because they are not only responsible for vaccinating themselves, but also their children. At the same time, recent outbreaks of preventable infectious diseases among children in the United States and Italy point to growing vaccination hesitancy among parents (Funk 2017; Nadeau 2015). Yet despite the importance of this group, there has been very little attention to factors influencing vaccination attitudes of parents. This lack of research is striking in that parents’ attitudes toward vaccinating not only impacts themselves, but also their children. To this end, our focus on vaccination attitudes of parents is theoretically appealing and possesses important implications for practice.

All participants were first asked to read a consent statement that explained the experiment and that there were zero risks involved in participating in the study. They were then asked to watch a short, 90-second video clip about a real outbreak of an infectious disease that occurred in the United States in 2015 for the sample of parents from the United States or in Italy in 2016 for the sample of Italian parents. The clip was included to enhance the credibility of the treatments and experimental setting. After watching the video clip, participants were randomly assigned to one of two groups. Both treatments explained that an outbreak of a dangerous infectious disease occurred in the United States (and Italy for Italian parents) and that a newly developed vaccine was available to members of the public for free. For the sample of US parents, treatments were written in English whereas for the sample of Italian parents, treatments were in Italian. To ensure language equivalence in treatments, the English treatment was translated into Italian and the Italian translation was back translated into English and compared with the original. For the statistical information group, numerical descriptors (e.g., $2,419.,325) were used to convey facts to participants. For the non-statistical information group, non-numeric descriptors (e.g., a couple million) were used. Additionally, the salience of statistical information has also been shown to vary according to the format the information is presented in (Porumbescu et al. 2017). This means that to ensure that effects we detect are not the result of choices on how to format our health information campaign and rather due to the statistical information itself we also randomly assigned participants to one of two blocks – one block broke content up into bullet point format, while the other present the content in block-paragraph format. Following exposure to treatments participants were then asked to respond to the same battery of survey items.

## Participants

Participants were 605 parents from the United States and 505 parents from Italy who were paid to participate in a survey response panels that work with the survey research firm Qualtrics. Participants were invited by Qualtrics to participate in our research project via email and could follow a link that would take them to the study. Quota sampling was used in order to ensure that the sample was representative of parents in the United States and Italy on the following dimensions: age, gender, and education, income, and political orientation. Characteristics of both samples are listed in tables 1a and 1b. A chi-square analysis of treatment groups revealed successful randomization in that none of the differences observed across the treatment groups in both countries was significant at the .05 percent level.

**Table 1a:** Demographic information for sample of US parents.

|  | Statistical Information | gender | age | educ | income |
| --- | --- | --- | --- | --- | --- |
| N | No | 301 | 301 | 301 | 301 |
|  | Yes | 303 | 302 | 303 | 303 |
| Mean | No | 1.50 | 33.1 | 4.56 | 5.33 |
|  | Yes | 1.51 | 32.8 | 4.40 | 5.11 |
| Std. error mean | No | 0.0289 | 0.462 | 0.119 | 0.169 |
|  | Yes | 0.0288 | 0.545 | 0.122 | 0.165 |

**Table 1b:** Demographic information for sample of Italian parents.

|  | Statistical Information | gender | age | educ | income |
| --- | --- | --- | --- | --- | --- |
| N | No | 259 | 259 | 259 | 259 |
|  | Yes | 248 | 248 | 248 | 248 |
| Mean | No | 1.53 | 36.2 | 3.44 | 3.31 |
|  | Yes | 1.45 | 43.9 | 3.40 | 3.15 |
| Std. error mean | No | 0.0311 | 0.540 | 0.116 | 0.136 |
|  | Yes | 0.0317 | 7.87 | 0.118 | 0.141 |

## Measures of Key Variables

### Comprehension

Our measure of comprehension taps into participants’ declarative memory, which refers to our ability to recall factual information related to a specific event (Eichenbaum 2000). Prior work in educational psychology (Blankenship and Dansereau 2000) and political science (Prior and Lupia 2008) assess this representation of comprehension by asking study participants to respond to a series of close-ended questions pertaining to different aspects of particular issue or information they have just read (Mangen, Walgermo, & Brønnick, 2013). Following this procedure, we measure comprehension by asking all participants assigned to a treatment group the same three multiple-choice questions related to the health information campaign treatment they were exposed to. Moreover, a recent technical report revealed that health information campaigns encouraging members of the public to vaccinate tend to focus on explaining how vaccinations work and the implications and risks of vaccinating (ECDC 2017). As such, questions relate to *how the vaccination works, or process comprehension (2 items)*, and *risks of being vaccinated, or risk comprehension (1 item)*. Items can be found in appendix 3.

#### Intentions to Vaccinate

We assess parents’ intentions to comply with government’s request that they vaccinate themselves against the virus they read about and we also assess their intentions to vaccinate their children against the same virus. Descriptive statistics for both items are provided in table 1a and 1b in the appendix 1.

To gauge participant intentions to comply with requests that they vaccinate themselves we use a real effort approach (Brüggen & Strobel, 2007). A common criticism of survey experiments is their lack external validity. For example, whether a manipulation that causes variation in stated intentions to comply with requests to vaccinate will cause the same type of variation in terms of the actual compliance behaviors is tenuous. In real effort experiments participants are assigned tasks that require an investment of resources, such as energy (Rosaz & Villeval, 2012). The idea is that asking participants to exert real effort, will offer a closer approximation to real life behavioral intentions than evaluating participants’ stated intentions. Based upon the real-effort framework, we evaluate participants’ intentions to comply with requests to vaccinate by asking them to sign up for times slots and dates to vaccinate – a total of 672 unique possible dates and times (i.e.; x days and y time slots per day). In brief, we explain to participants that there is a new and highly contagious strain of measles afflicting either the United States or Italy and that their local health clinic is providing free vaccinations. We then tell them that, to ensure access to the vaccine, government officials are asking adults to sign up for as many time slots as possible. While this is not the same as actually signing up for a vaccination, our measure does require greater mental (thinking about what dates and times might work) and physical (actually clicking potentially multiple dates) effort thereby making it a better approximation of real compliance behaviors than measures asking participants how likely they are to comply with the government’s request.

Furthermore, we are not only interested in parents’ intentions to comply themselves, but also their intentions to encourage their children comply with the government request as well. Accounting for parents’ intentions to vaccinate their children in addition to themselves is theoretically appealing because it speaks to the broader societal impacts of health information campaigns’ efforts to induce compliance.

To measure parents’ intentions to vaccinate their children we ask them: *How likely would you be to vaccinate your children to protect them from the risks associated with an outbreak such as this? (Response scale: 0 = definitely not, 100 = Definitely)*.

## Empirical Results

### Hypothesis 1

We begin by reviewing results of a one-way ANOVA, which examines differences in the three types of comprehension we assess across the treatment group exposed to statistical information and the group exposed to non-statistical information for both samples of parents (USA and Italy). In appendix 2 we provide supplementary analyses of hypothesis 1 for both samples using the Region of Practical Equivalence procedure, which uses a Bayesian estimation approach.

### US Sample

We find a significant difference between the two treatment groups on process comprehension (F(1, 602) = 7.40, p = 0.007, η_p_^2^ = 0.012). Out of a maximum of two correct answers, the mean level of process comprehension for those assigned to the statistical information condition was 0.977 (SE = 0.046), while for those assigned to the non-statistical information condition, the mean level of process comprehension was 1.156 (SE = 0.047). In summary, for US parents the mean level of comprehension of how the vaccine works was significantly lower for those assigned to the statistical information condition when compared to those assigned to the non-statistical information condition.

On the other hand for the second form of comprehension, *risk comprehension*, we find no statistically significant difference across treatment groups (F(1, 602) = 0.678, p = 0.411, η_p_^2^ = 0.001). Out of a maximum of one correct answer, the mean level of risk comprehension for those assigned to the statistical information condition was 0.327 (SE = 0.027), while for those assigned to the non-statistical information condition, the mean level of risk comprehension was 0.296 (SE = 0.026). Figure 1 illustrates these effects.

**Figure 1:**
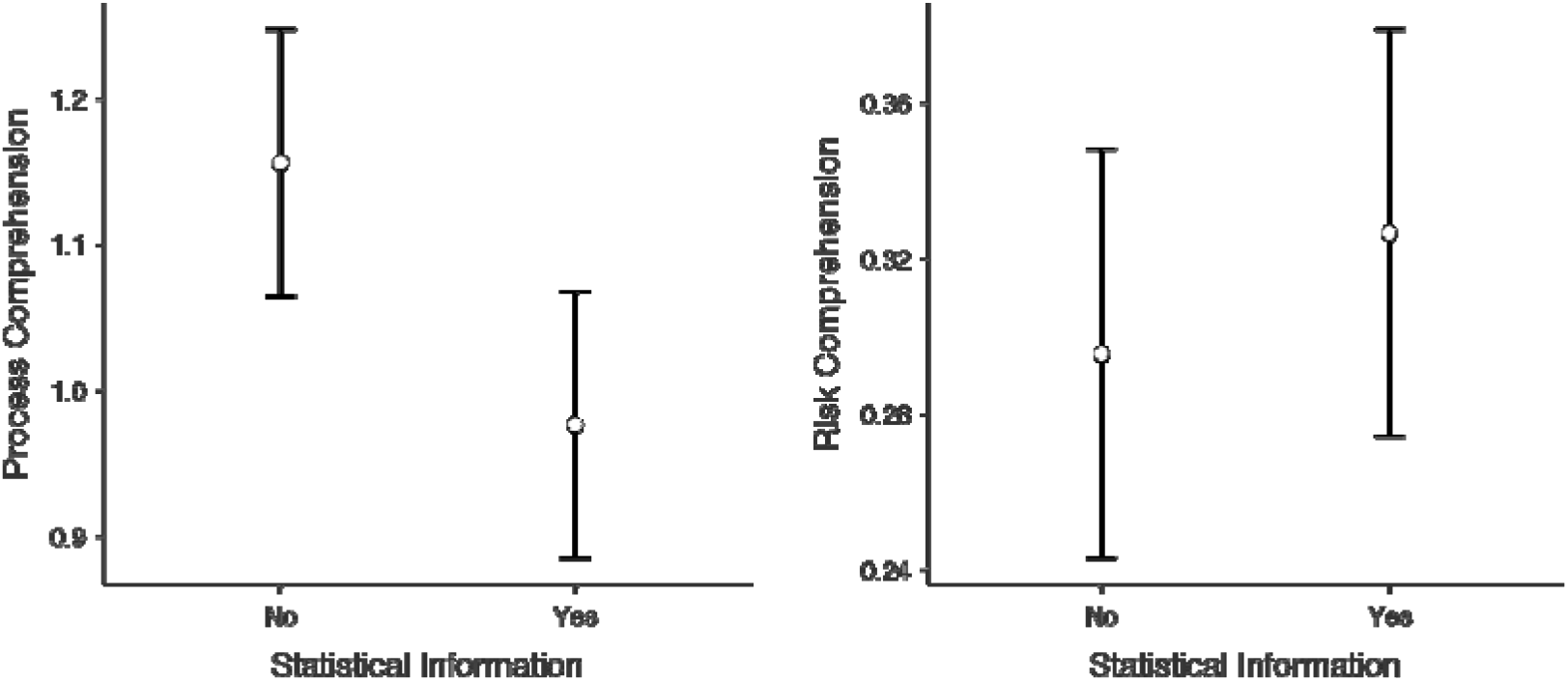
The Effect of Statistical Information on Process and Risk Comprehension for US Parents. Note: Error bars represent 95% confidence Intervals.

### Italian Sample

For the Italian sample we also find a statistically significant difference between the group exposed to statistical information and the group exposed to non-statistical information on process comprehension (F(1, 505) = 3.31, *p* = 0.070, η_p_^2^ = 0.007). However, as demonstrated by the level of statistical significance and effect size, this finding is not as strong as in the sample that uses US parents. Specifically, for parents assigned to the statistical information condition, the mean level of process comprehension was 1.29 (SE: 0.044) while for those exposed to the non-statistical information condition the mean level of process comprehension was 1.40 (SE: 0.043). p

Moreover, as with the sample of US parents, we also find no statistically significant difference across treatments groups on risk comprehension ((F(1, 505) = 1.92, *p* = 0.166, η_p_^2^ = 0.004). Out of a maximum of one correct answer, the mean level of risk comprehension for those assigned to the statistical information condition was 0.331 (SE = 0.029), while for those assigned to the non-statistical information condition, the mean level of risk comprehension was 0.274 (SE = 0.029).

**Figure 2:**
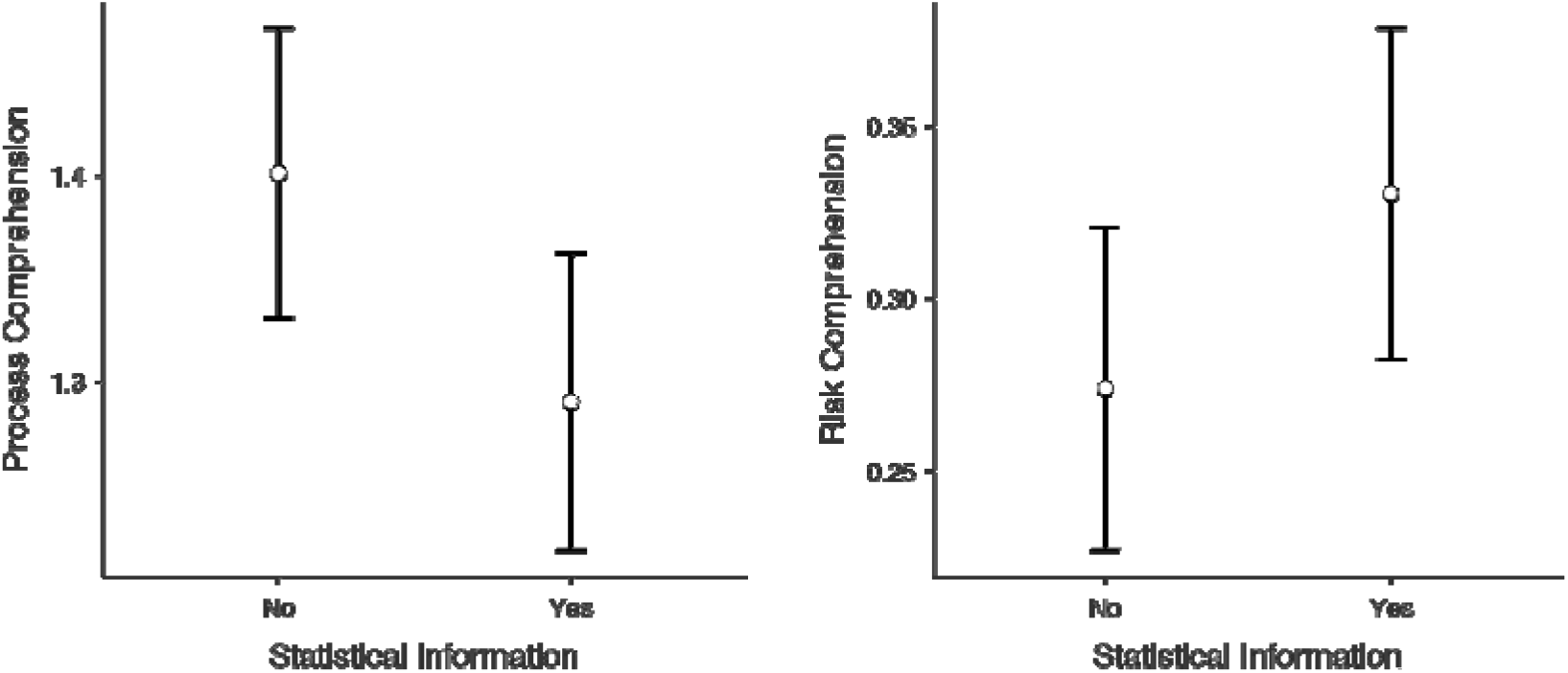
The Effect of Statistical Information on Process and Risk Comprehension for Italian Parents. Note: Error bars represent 90% confidence Intervals.

All told, we uncover a similar pattern of statistically significant and insignificant effects across the sample of parents from the United States and Italy. While hypotheses 1 predicted a general negative effect of statistical information on comprehension, these findings offer robust support of a more nuanced relationship between the inclusion of statistical information in health information campaigns and comprehension – statistical information matters more for process comprehension than risk comprehension. In the analysis to follow, we assess just how consequential process comprehension and risk comprehension are to compliance decisions. Figure 2 illustrates effects for both forms of comprehension.

### Hypothesis 2

Below we report findings on the relationship between two forms of comprehension and both measures of vaccination intentions through the use of ordinary least squares regression analysis.

### US Sample

Results point to a mixed relationship between different forms of comprehension and parents’ intentions to comply with government’s request that they vaccinate themselves. We find a positive association between process comprehension and parents’ intentions to comply with the request that they vaccinate themselves (*B* = 2.86, SE = 1.09, *p* = 0.009). That is, the better the parents understood how the vaccination worked, the more inclined they were to comply with the request vaccinate themselves. Moreover, we find that this relationship extends to parents’ intentions to vaccinate their children (*B* = 4.03, SE = 1.23, *p* = 0.001). As such, greater process comprehension was positively associated with parents’ intentions to comply with government’s request that they vaccinate themselves, but also extended to parents intentions to vaccinate their children.

Conversely, we see that participants’ risk comprehension was inversely related to their intentions to comply with the request to vaccinate themselves (*B* = –4.10, SE = 1.86, *p* = 0.027). That is, participants who understood the potential adverse effects of getting vaccinated expressed less of an interest in complying. This negative relationship is also present for parents’ intentions to vaccinate their children (*B* = –3.47, SE = 1.40, *p* = 0.013). In summary, parents who correctly answered the question about the possible adverse effects of vaccinating were less inclined to vaccinate themselves as well as their children.

Finally, we examine the interaction effect between the different forms of comprehension. That is, we look at how the relationship between process comprehension varies according to different levels of risk comprehension. Here were find that the relationship between process comprehension and parents’ intentions to vaccinate themselves does not significantly vary across levels of risk comprehension (*p* = 0.106). We do find a marginally significant interaction effect between the different forms of comprehension and parents’ intentions to vaccinate their children (*p* = 0.054). That is, the relationship between process comprehension and parents’ intentions to vaccinate their children was strongest among participants who incorrectly answered the question about the possible adverse effects of getting vaccinated. For ease of interpretation, we present a plot of this significant interaction effect in figure 3 below.

**Figure 3:**
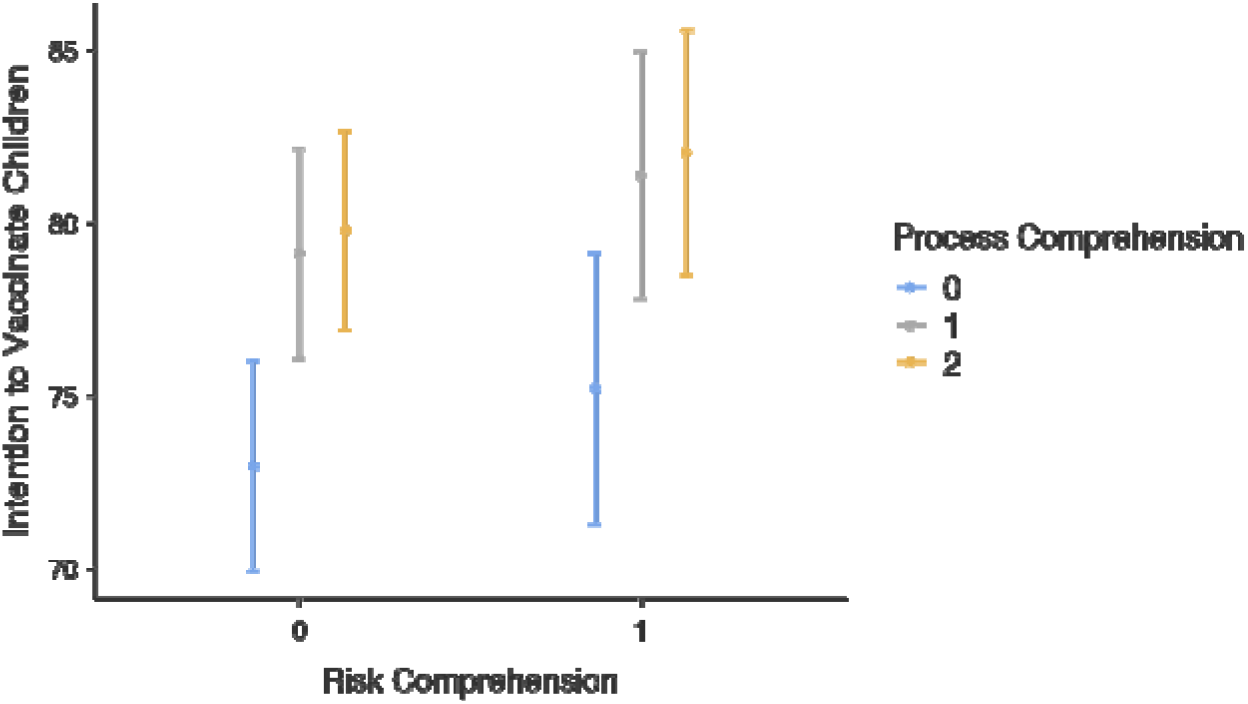
The relationship between Process comprehension and intentions to vaccination children at different levels of risk comprehension for US parents. Note: Error bars represent 90% confidence Intervals.

### Italian Sample

Results from the Italian sample point to a significant positive relationship between levels of process comprehension and parents’ intentions to comply with government’s request that they vaccinate themselves (*B* = 0.161, SE = 2.26, *p* < 0.000). The greater the comprehension of how the vaccine works, the greater the number of dates and times the Italian parents signed up for to get vaccinated. For the Italian parents’ intentions to vaccinate their children, we also find a significant positive relationship between levels of process comprehension and intentions to vaccinate their children (*B* = 0.152, SE = 1.33, *p* < 0.000). Just as with the sample of US parents, the sample of parents from Italy shows that higher levels of process comprehension is positively associated with parents intentions to vaccinate themselves and their children.

We find no statistically significant relationship between risk comprehension and Italian parents’ intentions to vaccinate themselves (*B* = –0.615, SE = 3.44, *p* = 0.858). In other words, understanding the potential negative impacts of vaccinating had little bearing on parents’ intentions to comply with the request to vaccinate themselves. We do however find a statistically significant and negative relationship between risk and Italian parents’ intentions to vaccinate their children (*B* = –3.73, SE = 2.01, *p* = 0.064). In sum, while understanding potential adverse effects of vaccinating had little bearing on parents’ intentions to comply with requests to vaccinate themselves, this form of comprehension was negatively related to parents’ intentions to vaccinate their children.

Finally, we find that the relationship between process comprehension and parents intentions to vaccinate themselves does not vary significantly according to levels of risk comprehension (*B* = –0.615, SE = 3.44, *p* = 0.858). However, just as with the sample of parents from the United States, we find that the relationship between process comprehension and Italian parents’ intentions to vaccinate their children did vary significantly according to risk comprehension (*B* = –3.73, SE = 2.01, *p* = 0.064). Specifically, as comprehension of the possible adverse effects of vaccinating increased, the association between comprehension of how the vaccine worked and parents’ intentions to vaccinate their children weakened. For ease of interpretation, we present a plot of this significant interaction effect in figure 4 below.

**Figure 4:**
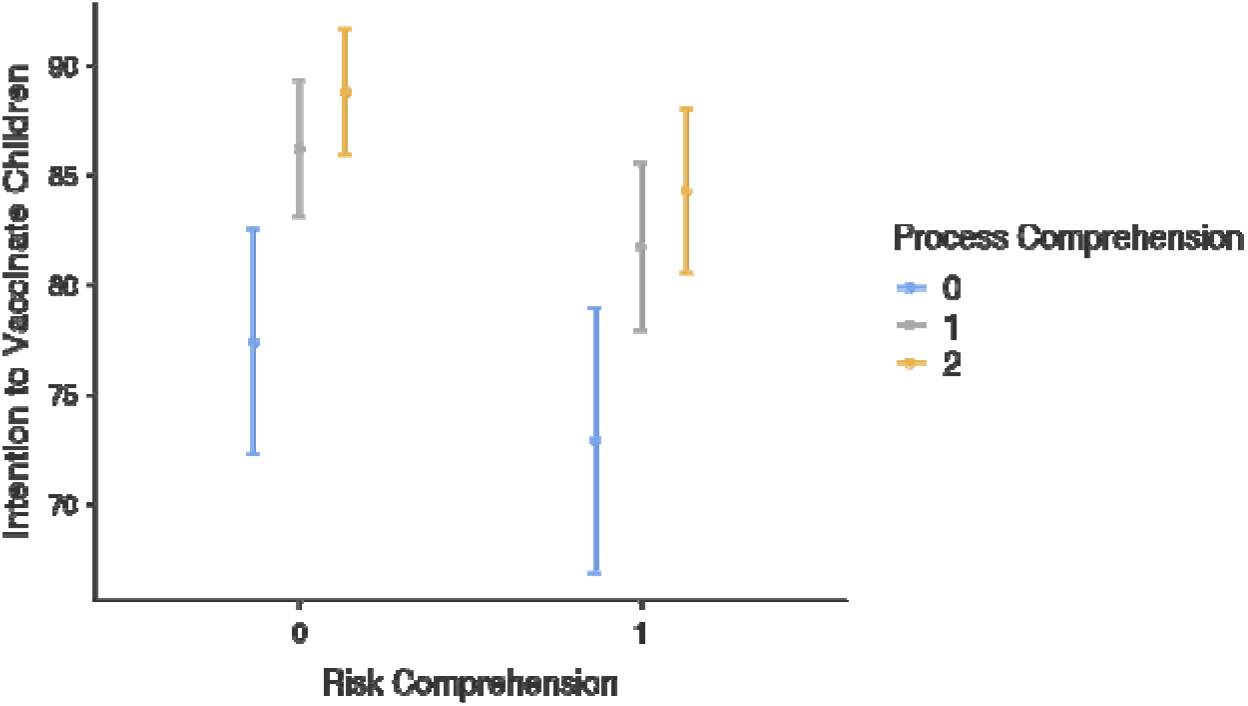
The relationship between Process comprehension and intentions to vaccination children at different levels of risk comprehension for US parents. Note: Error bars represent 90% confidence Intervals.

### Testing for indirect effects

Cumulatively, our hypotheses propose a sequence, such that statistical information will reduce comprehension of vaccination information, which in turn reduce intentions to vaccinate. Therefore, we test for indirect effects using causal meditational analysis proposed by Imai and colleagues (2011; 2013; see also Pearl 2012). This approach estimates indirect effects using a potential outcomes framework, which is advantageous when compared to the product of coefficients approach because the causal identification strategy is based upon counterfactual comparisons (Imai et al. 2013)^1^. Indirect effects calculated using this identification strategy are estimated using the ‘mediation’ R package (Tingley et al. 2014). We also evaluate the robustness of indirect effects by conducting a sensitivity analysis developed by Imai and colleagues (2011).

The results below detail indirect effects where our mediator is process comprehension. We do not test for indirect effects where comprehension of possible adverse effects acts as a mediator because there is no main effect of our treatment (statistical information) on this form of comprehension (see Hayes 2013).

### US Sample

We find evidence of a significant negative indirect effect of statistical information on parents’ intentions to comply with requests that they vaccinate themselves (*B* = – 0.490, CI 95 percent = –1.164, –0.06, *p* = 0.01). That is, our data indicate that statistical information decreases parents’ intentions to vaccinate themselves by suppressing participants’ process comprehension. Similarly, we find evidence of a significant negative indirect effect of statistical information on parents’ intentions to vaccinate their children (*B* = –0.651, CI 95 percent = –1.393, –0.12, *p* = 0.006). Sensitivity factors for both indirect effect estimates are 0.1. These findings indicate that including statistical information in our treatment suppressed process comprehension, which in turn reduced the likelihood of participants opting to comply with requests to vaccinate themselves, as well as their children.

### Italian Sample

We find evidence of a significant negative indirect effect of statistical information on parents’ intentions to comply with requests that they vaccinate themselves (*B* = – 1.063, CI 95 percent = –2.393, –0.03, *p* = 0.04). Just as in the sample of US parents we find that statistical information decreases parents’ intentions to vaccinate themselves by reducing their level of process comprehension. We also find evidence of a significant negative indirect effect of statistical information on parents’ intentions to vaccinate their children (*B* = –0.509, CI 95 percent = –1.331, –0.001, *p* = 0.048). Sensitivity factors for parents’ intentions to vaccinate themselves as well as their children were 0.2 indicating the indirect effects we identify using the sample of Italian parents are more robust than those uncovered in the sample of US parents.

All told, across both samples of parents, we find that the use of statistical information in health information campaigns reduces process comprehension and, in turn lowers intentions to comply with government requests that parents vaccinate themselves, as well as their children.

## Discussion and Conclusion

Health information campaigns are frequently used to elicit intentions to vaccinate, yet theoretical frameworks that guide their application are under-developed. In turn, this study draws upon the ELM CLT to develop and test an initial theoretical framework that offers insight into factors that shape the relationship between health information campaigns and intentions to vaccinate. Specifically, we argue that interventions that enhance comprehensibility of health information campaigns play a vital role in persuading citizens to vaccinate. In independent samples of parents from the United States and Italy we demonstrate that the presence of distractions in health information campaigns, namely including statistical information to explain facts to the public, negatively impacts a form of comprehension (i.e., process comprehension) that is critical to eliciting intentions to vaccinate. Before discussing contributions of these findings, we first outline limitations that pave the way for future research.

A first limitation of this study pertains to external validity. Specifically, the mechanisms we focus on address compliance intentions as they relate to a specific and controversial issue – vaccinations. We chose intentions to comply with vaccination requests to test the importance of comprehension as a mechanism because decisions to vaccinate are said to be accompanied by a strong emotional/irrational component, meaning that if comprehension is related to decisions to comply with requests to vaccinate it is likely to be related to many other forms of compliance, as well. This point however does not change that fact that conceptual replications, testing the relationships uncovered in this study as they relate to other forms of compliance is necessary. Second, in testing for indirect effects, we are unable to randomly assign participants to different levels of comprehension. In turn, this leads us to assume that there are no confounding variables biasing the relationship between process comprehension and vaccination intentions. While we find the presence of the indirect effect in very different settings (both samples of parents) where possible confounding variables would differ, caution should still be exercised in interpreting these results. A final point of caution concerns the small effect size of statistical information on process comprehension. While we find statistically significant effects of our treatment in both samples thus suggesting these effects are ‘real’, it is important to also note that using statistical information to convey facts is unlikely to induce dramatically different levels of process comprehension. That said, as our indirect effects reveal, the small impact that statistical information does have on comprehension does appear to carry important implications for decisions to comply with vaccination requests.

These limitations notwithstanding, our findings offer important contributions to the study of health information campaigns. First, this study contributes to the study of health information campaigns by developing a theoretical framework that deepens our understanding of how health information campaigns can be used to influence citizen intentions to vaccinate. This study does this by not only shedding light on factors that contribute to the success of these campaigns, but also by identifying a distinctive mechanism that helps to understand why some health information campaigns are more effective than others in triggering vaccination intentions.

A second contribution relates to discussions over strategies for effectively communicating government information to the public. While the use of statistical information is often thought to improve public service legibility (James 2011) and inspire greater confidence in the information source (Pederson 2017), as well as in the information itself (Zhang and Schwartz 2012), it can also serve as a distraction and inhibit information recall (Olsen 2017). Our findings offer support for prior work that finds the use of statistical information to convey facts to the public decreases information comprehension. Moreover, they also add to this discussion by showing how the use of statistical information can actually inhibit the persuasiveness of government communications by depressing forms of comprehension that are central to attitude change. As such, these findings are important important to the study of citizen vaccination intentions

Today, many of the health challenges confronting governments require flexible strategies that involve inputs from non-government actors, including members of the general public. With this in mind, health information campaigns can play a critical role in fostering collaboration with the general public to efficiently and effectively address social challenges, such as outbreaks of infectious diseases. However, the efficacy of these campaigns in eliciting cooperation from the general public is contested. Overall, we contribute to this discussion, demonstrating that health information campaigns can play an important role in fostering government collaboration with the public while also offering a framework for identifying interventions to enhance the effectiveness of these campaigns.

## Data Availability

Data are available upon request

## Appendix

## Appendix 1

**Table 1a:** Descriptive Statistics for Main Variables (US Parents)

|  | Group | Risk<br>Comprehension | Process<br>Comprehension | Intention<br>to<br>Vaccinate | Intention<br>to<br>Vaccinate<br>Children |
| --- | --- | --- | --- | --- | --- |
| N | No<br>statistical<br>information | 301 | 301 | 301 | 301 |
|  | Statistical<br>information | 303 | 303 | 303 | 303 |
| Mean | No<br>statistical<br>information | 0.296 | 1.16 | 79.7 | 76.9 |
|  | Statistical<br>information | 0.327 | 0.977 | 82.1 | 79.5 |
| Std.<br>error<br>mean | No<br>statistical<br>information | 0.0263 | 0.0473 | 1.38 | 1.43 |
|  | Statistical<br>information | 0.0270 | 0.0459 | 1.19 | 1.34 |

**Table 1b:** Descriptive Statistics for Main Variables (Italian Parents)

|  | Group | Risk<br>Comprehension | Process<br>Comprehension | Intention<br>to<br>Vaccinate | Intention to<br>Vaccinate<br>Children |
| --- | --- | --- | --- | --- | --- |
| N | No<br>statistical<br>information | 259 | 259 | 259 | 259 |
|  | Statistical<br>information | 248 | 248 | 248 | 248 |
| Mean | No<br>statistical<br>information | 0.274 | 1.40 | 83.1 | 85.3 |
|  | Statistical<br>information | 0.331 | 1.29 | 83.0 | 84.7 |
| Std.<br>error<br>mean | No<br>statistical<br>information | 0.0278 | 0.0421 | 1.32 | 1.29 |
|  | Statistical<br>information | 0.0299 | 0.0445 | 1.45 | 1.32 |

## Appendix 2: Results from Bayesian Parameter Estimation

Here we provide an alternative assessment of the strength of evidence we find related hypothesis 1 that is based upon a Bayesian estimation of parameters. Following Krushke (2018) this approach to equivalence testing has two key elements: 1) the 95% Highest Density Interval (HDI), which reflects 95% of the most credible parameter estimates, and 2) the region of practical equivalence (ROPE), which identifies the credible parameter values that are practically equivalent to the null. Using the HDI + ROPE framework developed by Krushke (2018), we assess what percentage of credible parameter estimates fall within the ROPE. The lower the percentage of parameter estimates that fall within the ROPE the stronger our evidence in support of hypothesis 1.

### US Parents

**Figure A1:**
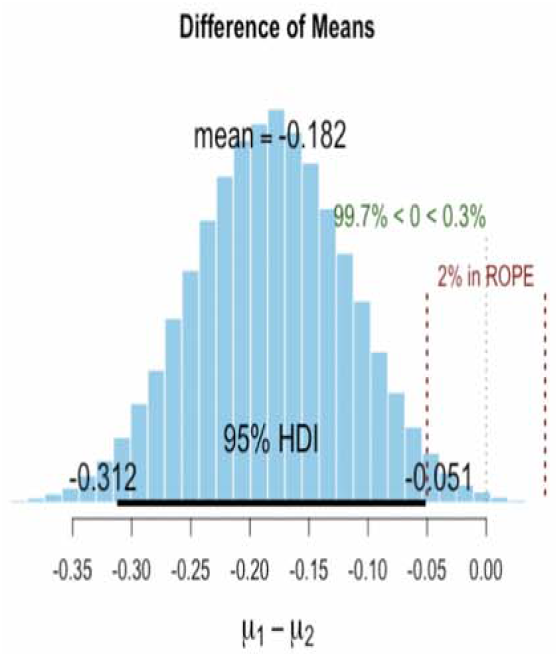
Posterior Distribution of Difference of Means for <u>Process Comprehension</u>. Note: Bounds on the ROPE reflect ±5% in process comprehension. These results correspor to the marginally significant effect identifiedfor the sample of US parents.

**Figure A2:**
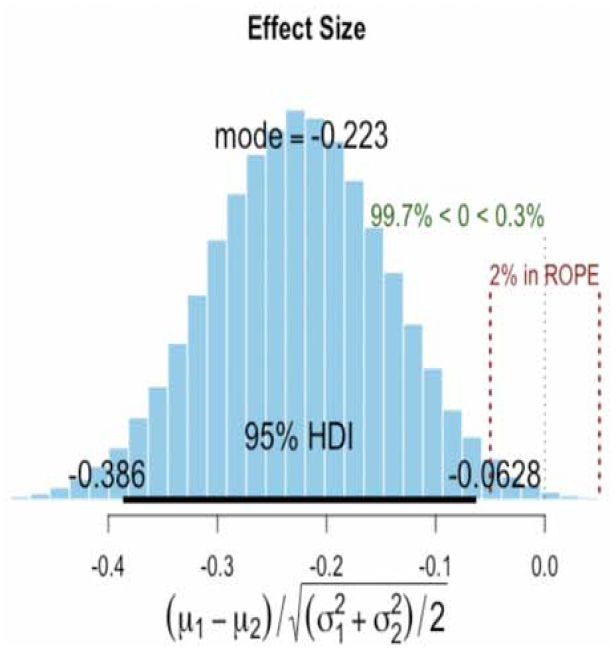
Posterior Distribution of Effect Size cf Statistical Information on <u>Process Comprehension</u>. Note: Bounds on the ROPE reflect ±5% one is able to achieveone is able to achievein process comprehension. These results correspond to the marginally significant effect identified for the sample of US Parents.

**Figure A3:**
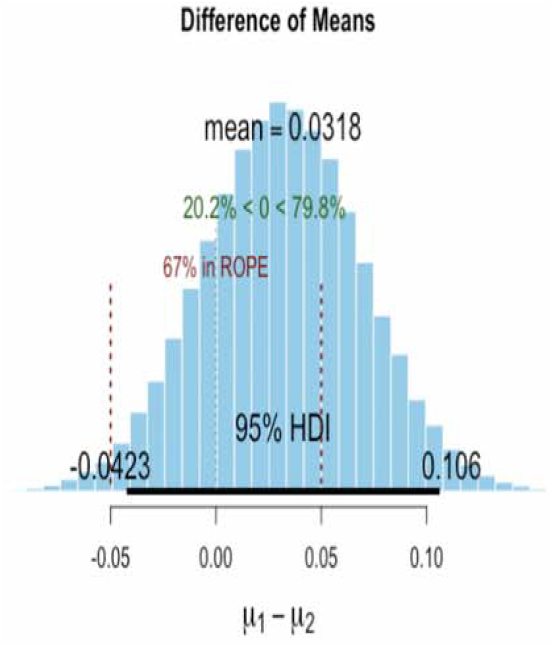
Posterior Distribution of Difference of Means for <u>Risk Comprehension</u>. Note: Bounds on the ROPE reflect ±5% in process comprehension. These results correspon, to the marginally significant effect identified for the sample of US parents.

**Figure A4:**
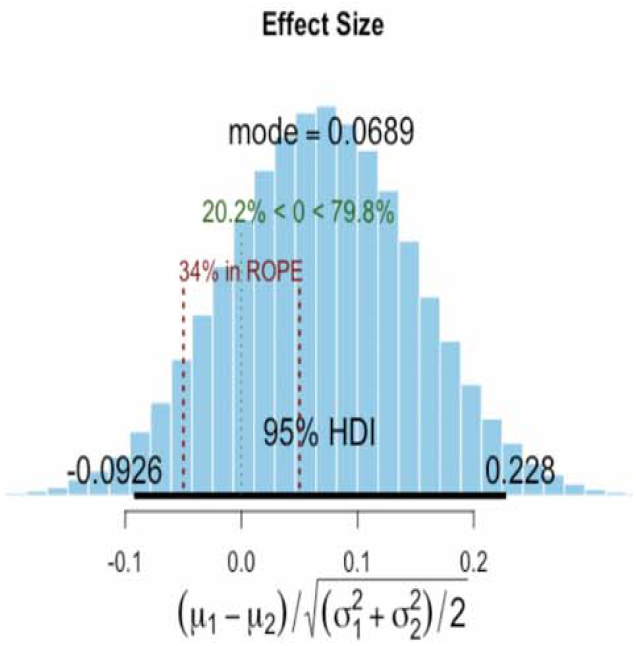
Posterior Distribution of Effect Size of Statistical Information on <u>Risk Comprehension</u>. Note: Bounds on the ROPE reflect ±5% in process comprehension. These results correspond to the marginally significant effect identified for the sample of US parents.

### Italian Parents

**Figure A5:**
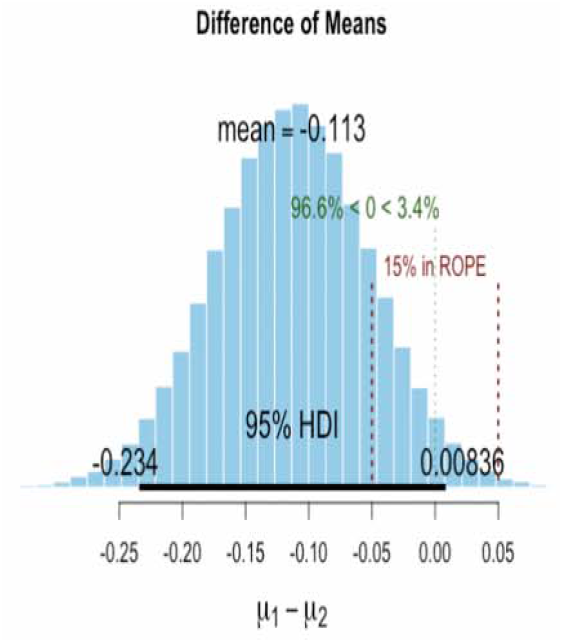
Posterior Distribution of Difference of Means for <u>Process Comprehension</u>. Note: Bounds on the ROPE reflect ±5% in process comprehension. These results correspond to the marginally significant effect identified for the sample of Italian Parents.

**Figure A6:**
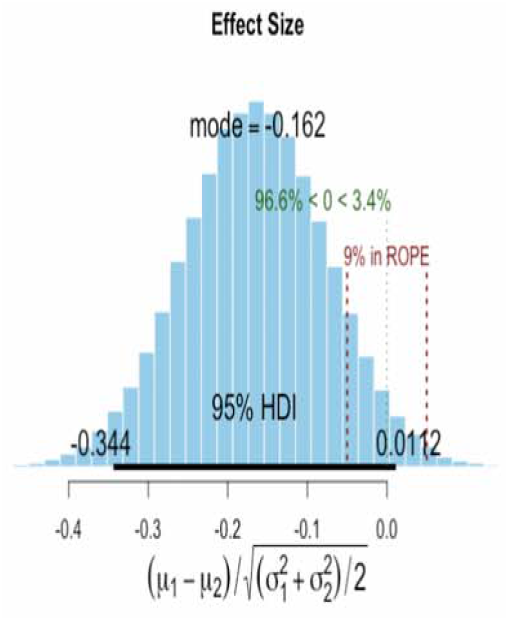
Posterior Distribution of Effect Size of Statistical Information on <u>Process Comprehension</u>. Note: Bounds on the ROPE reflect ±5% in process comprehension. These results correspond to the marginally significant effect identified for the sample of Italian Parents.

**Figure A7:**
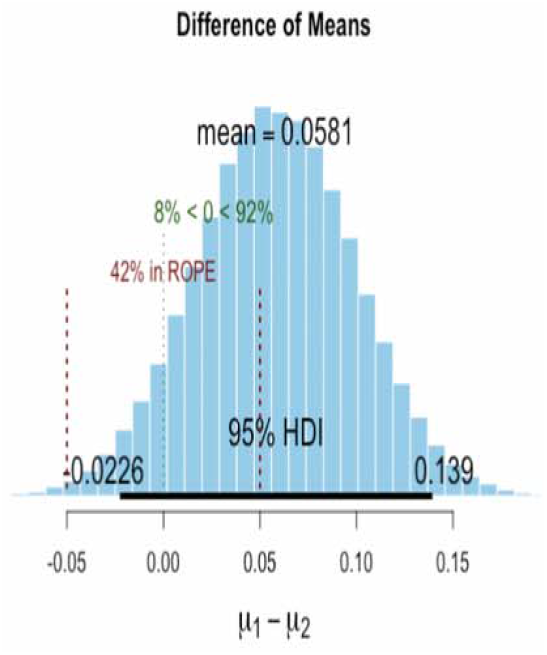
Posterior Distribution of Difference of Means for <u>Risk Comprehension</u>. Note: Bounds on the ROPE reflect ±5% in process comprehension. These results correspond to the marginally significant effect identified for the sample of Italian parents.

**Figure A8:**
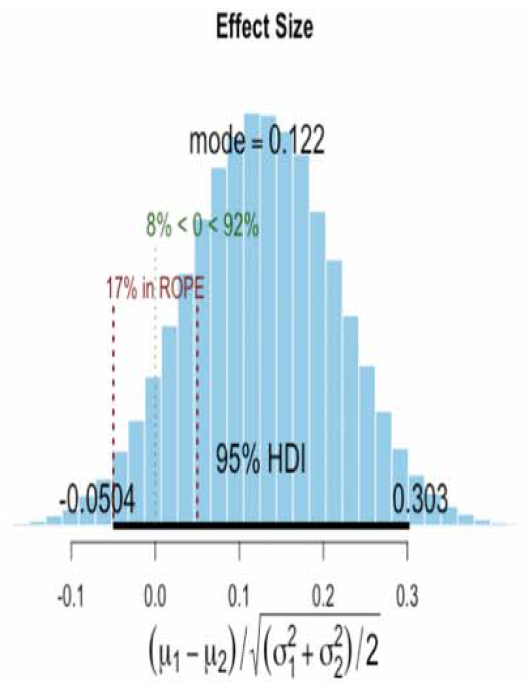
Posterior Distribution of Effect Size of Statistical Information on <u>Risk Comprehension</u>. Note: Bounds on the ROPE reflect ±5% in process comprehension. These results correspond

## Appendix 3: Materials

### Statistical Information Treatment X Block Paragraph Text

83% of the United States population will be affected by an outbreak of a severe and highly infectious new form of measles. Studies show that 67.1% of those infected will die. 97.5% of those infected are expected to suffer major health issues as a result. Scientists at The Center for Disease Control recently spent $2,419,325 to develop a vaccine that can protect citizens from this virus. The vaccine works by injecting 0.00003 CCs of the potentially deadly virus into a healthy person and then letting their body develop immunity. Among those who receive the vaccine 2.4% are expected to experience complications, whereas for 0.4% the vaccination may prove fatal. The vaccination will be provided to those who want it for free.

### No Statistical Information Treatment X Block Paragraph Text

The majority of the United States population will be affected by an outbreak of a severe and highly infectious new form of measles. Studies show that around 2/3 of those infected will die. Almost all of those infected are expected to suffer major health issues as a result. Scientists at The Center for Disease Control recently spent a couple million dollars to develop a vaccine to protect citizens from this virus. The vaccine works by injecting a very small quantity of the deadly virus into a healthy person and then letting their body develop immunity. Among those who receive the vaccine a very small number are expected to experience complications, whereas fatalities due to the vaccine will be extremely rare. The vaccination will be provided to those who want it for free.

### Statistical Information Treatment X Bulleted Text

- 83% of the United States population will be affected by an outbreak of a severe and highly infectious new form of measles.
- Studies show that 67.1% of those infected will die. 97.5% of those infected are expected to suffer major health issues as a result.
- Scientists at The Center for Disease Control recently spent $2,419,325 to develop a vaccine that can protect citizens from this virus.
- The vaccine works by injecting 0.00003 CCs of the potentially deadly virus into a healthy person and then letting their body develop immunity.
- Among those who receive the vaccine 2.4% are expected to experience complications.
- For 0.4% of the population the vaccination may prove fatal.
- The vaccination will be provided to those who want it for free.

### No Statistical Information Treatment X Bulleted Text

- The majority of the United States population will be affected by an outbreak of a severe and highly infectious new form of measles.
- Studies show that more than half of those infected will die.
- Almost all of those infected are expected to suffer major health issues as a result.
- Scientists at The Center for Disease Control recently spent a couple million dollars to develop a vaccine to protect citizens from this virus.
- The vaccine works by injecting a very small quantity of the deadly virus into a healthy person and then letting their body develop immunity.
- Among those who receive the vaccine a very small number are expected to experience complications.
- Fatalities due to the vaccine will be extremely rare.

The Vaccination will be provided for free to those who want it.

### Measures of comprehension

#### Risk Comprehension (1 Item)

For those who get vaccinated

- There is no chance of complications and no risk of dying (1)
- There is a small chance of complications, but no risk of dying (2)
- There are no documented complications, but a small risk of death (3)
- There is a small chance of complications, and very small chance of dying (4)
- We don’t know whether there are side effects (either complications or deaths) as there have been no attempts to examine this (5)

#### Process Comprehension (2 Items)

1. Scientists have
  - Not had enough time to develop an effective vaccine for the outbreak (1)
  - Developed a vaccine that can protect individuals from the outbreak (2)
  - Limited supplies of an effective vaccine to protect individuals from the outbreak (3)
  - Developed a partially effective vaccine to protect individuals from the outbreak (4)
  - Warned that there is no vaccine that can be developed to stop the outbreak (5)
2. How does the vaccine work?
  - A large quantity of the deadly virus is injected into your body (1)
  - A miniscule quantity of the deadly virus is injected into your body (2)
  - A large quantity of free radical fighting anti-oxidants are injected into your body (3)
  - The prompt did not explain (4)

## Footnotes

1 Under the potential outcomes framework, an indirect effect is estimated using the following structural equation: Yi = α_i_ + β_i_ T_i_+ γM_i_ + ε_i_. Using binary treatment and mediators for illustrative purposes, we first predict an outcome value, Y, which results from the value of the mediator calculated under treatment (m(t_1_)). Next, calculate the outcome under treatment, but using a value of the mediator that corresponds to the control condition (m(t_0_)). The Average causal mediated effect corresponds to the average difference between these two. In other words, our indirect effect is: Y_1_(t_1_, m(t_1_) – Y_2_(t_1_, m(t_0_).

